## Supplementary Information and Tables for "Machine learning assisted analysis on TCR profiling data from COVID-19-convalescent and healthy individuals unveils cross-reactivity between SARS-CoV-2 and a wide spectrum of pathogens and other diseases": Supplementary_Information.pdf

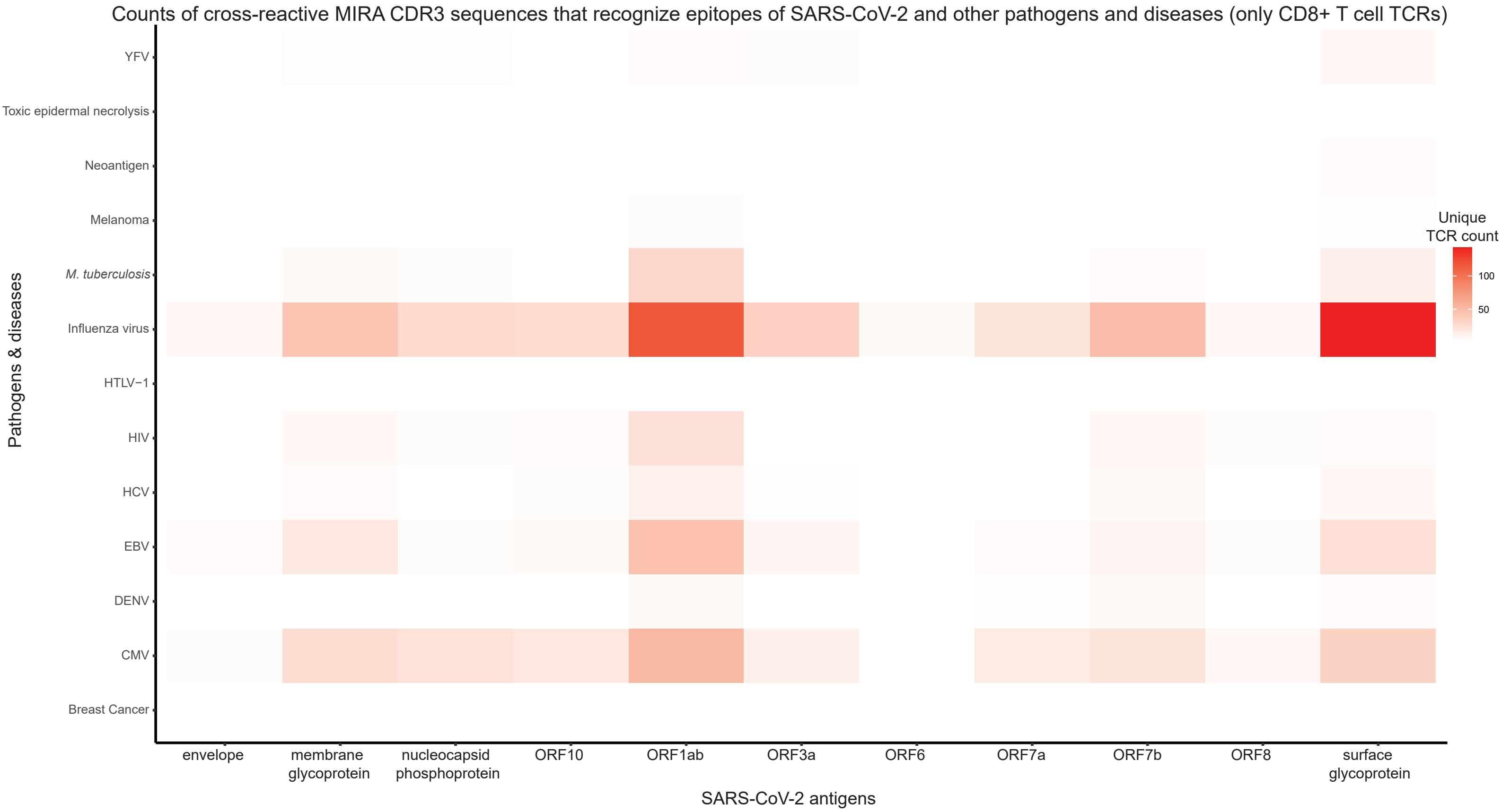

Counts of cross-reactive MIRA CDR3 sequences that recognize epitopes of SARS-CoV-2 and other pathogens and diseases (only CD8+ T cell TCRs)
Pathogens & diseases
YFV
Toxic epidermal necrolysis
Neoantigen
Melanoma
M. tuberculosis
Influenza virus
HTLV−1
HIV
HCV
EBV
DENV
CMV
Breast Cancer
SARS-CoV-2 antigens
envelope
membrane glycoprotein
nucleocapsid phosphoprotein
ORF10
ORF1ab
ORF3a
ORF6
ORF7a
ORF7b
ORF8
surface glycoprotein
Unique TCR count
100
50

Supplementary Figure 1. Cross-reactivity analysis of all Multiplexed Identification of T cell Receptor Antigen (MIRA) T cell receptors (TCRs) only from CD8+ T cells. The cross-reactivity is depicted as a heatmap of unique MIRA complementarity-determining region 3 (CDR3) counts that exhibit cross-reactivity between SARS-CoV-2 (x-axis) and other pathogens and diseases (y-axis).
